# Estimating internationally imported cases during the early COVID-19 pandemic

**DOI:** 10.1101/2020.03.23.20038331

**Authors:** Tigist F. Menkir, Taylor Chin, James Hay, Erik D. Surface, Pablo M. De Salazar, Caroline O. Buckee, Alexander Watts, Kamran Khan, Ryan Sherbo, Ada W. C. Yan, Michael Mina, Marc Lipsitch, Rene Niehus

**Affiliations:** Center for Communicable Disease Dynamics, Department of Epidemiology, Harvard T.H. Chan School of Public Health, Harvard University, Boston, Massachusetts, USA; Li Ka Shing Knowledge Institute, St. Michael’s Hospital, Toronto, Canada; Department of Medicine, Division of Infectious Diseases, University of Toronto, Canada; BlueDot, Toronto, Canada; Section of Immunology of Infection, Department of Infectious Disease, Imperial College London, London, United Kingdom

**Keywords:** outbreak, Wuhan, China, underdetection, prevalence, COVID-19, flight data, travel ban, Africa

## Abstract

Early in the COVID-19 pandemic, when cases were predominantly reported in the city of Wuhan, China, local outbreaks in Europe, North America, and Asia were largely predicted from imported cases on flights from Wuhan, potentially missing imports from other key source cities. Here, we account for importations from Wuhan and from other cities in China, combining COVID-19 prevalence estimates in 18 Chinese cities with estimates of flight passenger volume to predict for each day between early December 2019 to late February 2020 the number of cases exported from China. We predict that the main source of global case importation in early January was Wuhan, but due to the Wuhan lockdown and the rapid spread of the virus, the main source of case importation from mid February became Chinese cities outside of Wuhan. For destinations in Africa in particular, non-Wuhan cities were an important source of case imports (1 case from those cities for each case from Wuhan, range of model scenarios: 0.1-9.8). Our model predicts that 18.4 (8.5 - 100) COVID-19 cases were imported to 26 destination countries in Africa, with most of them (90%) predicted to have arrived between 7th January (±10 days) and 5th February (±3 days), and all of them predicted prior to the first case detections. We finally observed marked heterogeneities in expected imported cases across those locations. Our estimates shed light on shifting sources and local risks of case importation which can help focus surveillance efforts and guide public health policy during the final stages of the pandemic. We further provide a time window for the seeding of local epidemics in African locations, a key parameter for estimating expected outbreak size and burden on local health care systems and societies, that has yet to be defined in these locations.

## Introduction

In late December 2019, a new severe acute respiratory syndrome coronavirus 2 (SARS-CoV-2) was identified, in Wuhan, Hubei Province, China.^1^ Rigorous measures to curtail the spread of SARS-CoV-2, the causative virus of the COVID-19 disease, including travel restrictions and school and workplace closures, have largely controlled the outbreak in mainland China.^2–5^ However, international exportation of COVID-19 cases before the outbreak was contained in mainland China ignited global spread of COVID-19,^6^ which has now become a pandemic. As of 6th July 2020, 1.1 million confirmed cases of COVID-19 have been registered worldwide, with 85 thousand detected in mainland China and the remainder detected internationally in 209 other locations.^7^

Because the majority of cases in the early phase of the pandemic were reported in Wuhan, early COVID-19 case definitions and clinical guidelines required individuals suspected of infection to have had a recent travel history from Wuhan.^8^ On a similar vein, models predicting internationally imported cases from China and local outbreaks in North America, Europe and Asia have largely relied on flight passenger numbers from Wuhan.^6,9^ Based on those models, the risk for outbreaks in several African countries was estimated to be relatively low.^9^ However, a significant number of COVID-19 cases were introduced to other large cities in China before travel restrictions were instituted on 23rd January 2020.^10,11^ This suggests that there may have been a substantial risk of early case exportation from Chinese cities outside of Wuhan, that could have led to additional undetected imported cases globally, and to an elevated risk of importation to African countries specifically.

As of 6th July 2020, more than 478,000 confirmed cases have been reported in 55 African locations.^7^ Coverage of COVID-19 diagnostic and control interventions in many of those locations is expanding yet still limited, and many of these nations will continue to struggle to meet increased demand.^12–14^ Furthermore, in the absence of reliable estimates of prevalence, it is extremely difficult to assess the current and future burden of COVID-19 in many of these locations. In light of these challenges, accurately estimating the timing and number of initial importations is crucial to inform models of outbreak dynamics for those locations.

Previous work has combined travel data with incidence estimates to estimate the risk of importation from all Chinese provinces, excluding Hubei, to all African countries.^15^ That analysis, however, used historical flight data from January 2019, which is unlikely to reflect 2020 travel trends, given that the Lunar New Year occurred comparatively early (in January) this year and unprecedented travel restrictions were in place starting late January. In addition, the analysis did not take into account the uncertainty in reporting rates, the delay between infection and case report, and the time-varying prevalence of infected individuals in China.

Here, we have developed a modelling framework to synthesize available travel and COVID-19 prevalence data to explore geographical and time-varying trends of international case importation. We first estimated daily flight passenger numbers from 18 major cities in China to 43 international destinations from December 2019 to February 2020. Importantly, we used current data on air travel that takes into account increased travel in early January due to the Lunar New Year holiday and reduced travel in late January due to travel restrictions. We then estimated daily COVID-19 prevalence in each Chinese province, taking into account delays between infection, symptom onset and confirmation, and differences in ascertainment rates between Hubei and other Chinese provinces. Finally, we combined air-travel and prevalence estimates to give the daily number of internationally imported cases from mainland China to 43 international locations, including 26 destinations in Africa and globally representative locations on every continent. A number of assumptions are required for our predictions (e.g. relative ascertainment rates between Hubei and other provinces and the duration that each infection contributes to prevalence), and we therefore present results from a range of scenarios capturing uncertainty in our key assumptions, with point estimates from our selected best-estimate scenario. Our findings reveal a shift in the importance of Wuhan versus other Chinese cities as a source for COVID-19 importations over the course of the outbreak, and a generally higher importance of non-Wuhan cities for case importations to African locations. Further, our model estimates the time window in which local outbreaks may have been initiated and signals heterogeneities in the extent and the exact source of introduced cases in the African locations.

## Results

### 1. Estimating the number of airplane passengers from China to global destinations

We estimated the daily air-travel volume, defined as the daily number of passengers on direct and indirect air-travel routes, from 18 different Chinese cities to 43 international destinations. We estimated this within our focal time period, between 1st December 2019, (the approximate date of initiation of the pandemic in China^16^), and 29th February 2020, as the most recent date for which we have flight information (Figure S1). The 18 Chinese origin cities have been previously identified as high-risk cities for importation of COVID-19 from Wuhan.^11^ Our 43 destinations included 1) ten *high-surveillance locations* that have high surveillance capacity index (discussed previously^6^) and high air-travel connectivity to Wuhan, 2) 26 destinations in Africa (*African locations*) with high air-travel connectivity to the 18 Chinese origin cities and finally 3) seven additional locations that together with a subset of 1) and 2) yield 16 *globally representative locations*, which receive worldwide the highest air-travel volume from China and also represent every inhabited continent. Air-travel volume on each day of our focal period for each origin-destination pair was calculated using monthly air-travel volume (number of flight passengers), which we apportioned into days using daily flight departures (number of departing flights, see Methods).

The resulting trends in daily air-travel volume reflect the timing of the Lunar New Year holiday in 2020, which occurred much earlier than in recent years, and also the impact of widespread travel restrictions in late January 2020 (Figure S1). Air-travel volume increased in January during the Chunyun holiday – the 40-day holiday period that began on 10th January 2020 and surrounds Lunar New Year – relative to the same time period in 2019 (Figure S1). The sharp decline in air-travel volume after 23rd January 2020 is a result of the travel restrictions and flight cancellations that occurred starting late January (Figure S1).

### 2. Estimating daily prevalence of COVID-19 in 18 Chinese cities

Next, we estimated a daily prevalence indicator for all Chinese cities considered in our analysis. We defined this prevalence indicator as a measure that is linearly proportional to the actual (unobserved) prevalence of infected cases that are able to travel. See Methods for a detailed description. In brief, we first estimated the province-level daily incidence of onsets for detected infections by shifting confirmed case count curves using the delay between infection and symptoms (incubation period) and the delay in reporting (Figure 1A, Figure S2). We then adjusted our estimates of incidence in Hubei to account for lower ascertainment of cases in Hubei relative to other provinces in China, in order to yield a measure of relative incidence in each province. Next, we estimated relative prevalent cases from incidence by assuming that each newly infected case contributed to province-level prevalence for a number of days before they were no longer included in the travel-relevant pool of infected individuals (Figure 1B) (e.g. due to quarantine upon symptom onset). Finally, we allocated those province-level prevalence indicators to each city included in our analysis and standardized by population size to compute the corresponding city-level prevalence indicators (Figure S3).

**Figure 1.**
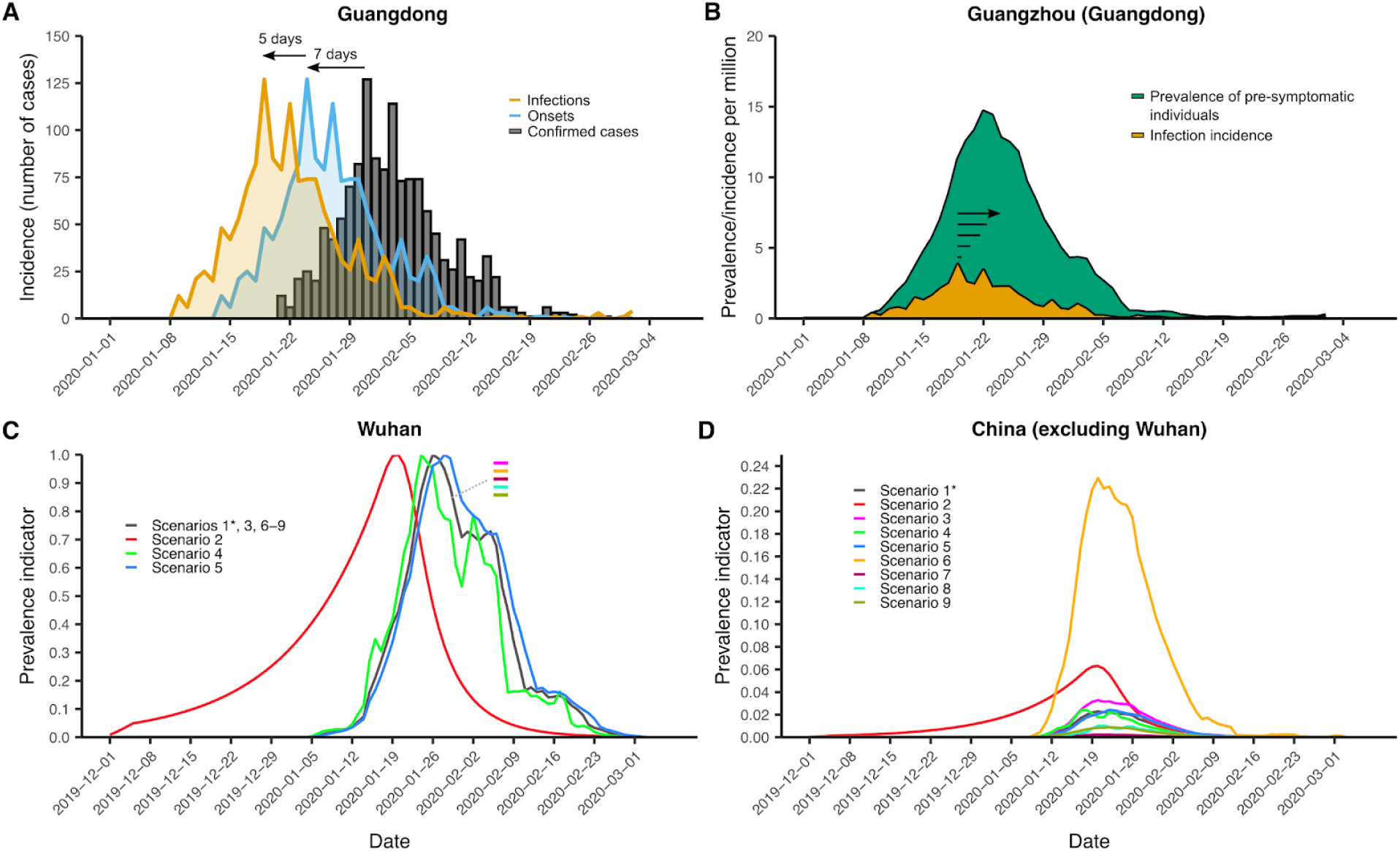
**(A)** Back-shifting of symptom onsets (blue) and infections (orange) from observed confirmed case counts (grey bars) using Guangdong as an example. Confirmed cases were shifted back by 7 days (the mean confirmation delay) to estimate symptom onset incidence, then further by 5 days (the median incubation period) to estimate infection incidence of those cases. **(B)** Conversion of province-level infection incidence to city-level pre-symptomatic prevalence in Guangzhou. Each infected individual (orange) was assumed to contribute towards pre-symptomatic prevalence (green) for 5 days (the median incubation period). Incidence and prevalence are shown per capita and before adjusting for within China differences in ascertainment rates. **(C)** and **(D)** Prevalence indicator for Wuhan and averaged for non-Wuhan cities under the 9 tested scenarios. The best-estimate scenario is Scenario 1 (highlighted with an asterisk). Note that for curves of this indicator only relative comparisons are meaningful, and thus they are scaled such that the highest peak is set to 1 within each scenario. Note further that in **(C)** Scenarios 1, 3 and 6-9 yield identical curves for Wuhan and that Scenario 2 is based on independent modeling estimates. Scenarios 1, 3, 6 and 7 only differ in their assumed ascertainment rate ratio, which we account for in all scenarios by scaling up (or in the case of Scenario 6, scaling down) incidence in Hubei and keeping constant incidence in all other cities.

In our best-estimate scenario (Scenario 1), we assumed that 1) each new infected case may travel for up to five days before showing symptoms (i.e. the median incubation period^17^), that 2) case ascertainment is twice as high outside of Hubei as it is in Hubei (based on data from Verity et al.^18^), and that 3) all cases reported in a province came from the city/cities included in our analysis, since larger cities considered to be ‘travel hubs’ are more likely to be the key contributor of infected cases. Under these assumptions, we found that the prevalence indicator peaked in all Chinese cities between 19th and 26th January 2020 (Figure 1 C&D and Figure S3). Other than Wuhan, Chinese cities with the highest prevalence indicators were Jiaxing, Nanchang and Changsha, whereas Beijing, Shanghai and Tianjin had the lowest.

To account for the considerable uncertainty in the true time-varying SARS-CoV-2 infection prevalence in Wuhan and elsewhere in China during the initial outbreak, we considered eight alternative scenarios to assess the impact of these assumptions on our subsequent analyses of case importations (Table 1, Figure 1C&D, Figure S3). A key alternative scenario used estimated incidence curves from a previous analysis that accounted for time-varying ascertainment rates in China due to changing case definitions (Scenario 2).^19^ In contrast to our best-estimate scenario, which shifts and inflates only observed case counts, this alternative estimate suggested substantial undocumented incidence throughout December and early January (Figure 1C&D). Note that the prevalence indicators from scenario 2 provide a measure of absolute, rather than relative, prevalence, as in all other scenarios. Under Scenario 2, prevalence indicators peaked on 20th January 2020 in all locations.

**Table 1.** Key parameter assumptions that we vary under the nine scenarios.

| Parameter | Scenario 1* | Scenario 2 | Scenario 3 | Scenario 4 | Scenario 5 | Scenario 6 | Scenario 7 | Scenario 8 | Scenario 9 |
| --- | --- | --- | --- | --- | --- | --- | --- | --- | --- |
| Ascertainment rate ratio | 2:1 | Time-varying | 1.4:1 | 2:1 | 2:1 | 0.2:1 | 20:1 | 2:1 | 2:1 |
| Days contributing to relative prevalent cases | 5 days | 5 days | 5 days | 2 days | 7 days | 5 days | 5 days | 5 days (Wuhan) and 2 days, following 3 day gap (outside of Wuhan) | 5 days |
| Allocation of relative prevalent cases to cities | Cities used assumed to capture all infections | Cities used assumed to capture all infections | Cities used assumed to capture all infections | Cities used assumed to capture all infections | Cities used assumed to capture all infections | Cities used assumed to capture all infections | Cities used assumed to capture all infections | Cities used assumed to capture all infections | Cities capture a fraction of infections equal to the proportion of the province population in that city |

\*Scenario considered most plausible (best-estimate scenario)

### 3. Predicting exported case counts to African countries

We combined our estimates of the daily COVID-19 prevalence indicator in China with the daily air-travel volume between China and a given set of destinations to obtain an indicator that is linearly proportional to the daily flight volume of infected travelers for each origin-destination pair, which we term the “force of importation.” To translate the force of importation into the expected actual number of imported cases to a destination, we extended on the approach outlined in De Salazar *et al*. (2020)^9^ and fit a Poisson regression model to the cumulative number of imported and detected cases (with identified source location Wuhan) in our *high-surveillance locations* from 1st December 2019 to 23rd January 2020, when the lockdown was instituted in the city. For this we adjusted each destination country’s detected case count by an estimate of underreporting^6^ and focused only on imported cases from Wuhan, as we assume that a majority of imported cases during this period of the pandemic originated from Wuhan. We use the Poisson model fit for high-surveillance locations, which captures the extent to which our estimated forces of importation must be scaled to align with the number of imported cases, to make predictions for other locations. Here, uncertainty around the nine scenarios for the prevalence indicator described previously (model uncertainty) far exceeded that from fitting the Poisson regression (statistical uncertainty), so we chose only to present any variation in our results across the nine scenarios, as this reflects the greatest uncertainty in our estimates. We bound our point estimates, i.e. mean predictions from Scenario 1, with the range in mean predictions across all nine model scenarios.

Our model estimates in our African destinations a total of 18.4 (8.5 - 100) imported COVID-19 cases, with approximately 100% predicted on days prior to the first case detections in each location. The highest numbers of imports are expected for South Africa (4.5; 1.7 - 18) and Egypt (3.9; 1.0 - 33), followed by Kenya (2.0; 0.8 - 9.2) and Algeria (1.6; 0.2 - 2.2). We estimate the lowest expected case counts in Equatorial Guinea and Mauritania (both 0.04; 0 - 0.2) (Figure 2). Figure 3A depicts the weekly predicted number of imported cases in each of the 26 African countries over time, highlighting the top five countries for which we predict the highest force of importation. All countries exhibit similar trajectories of predicted imported cases over the focal time period, with the exception of Algeria, for which our estimates suggest a slightly delayed initial increase in cases relative to the other top ranking locations. 90% of all imported cases were estimated to be imported between 7th January (±10 days) and 5th February (±3 days).

**Figure 2.**
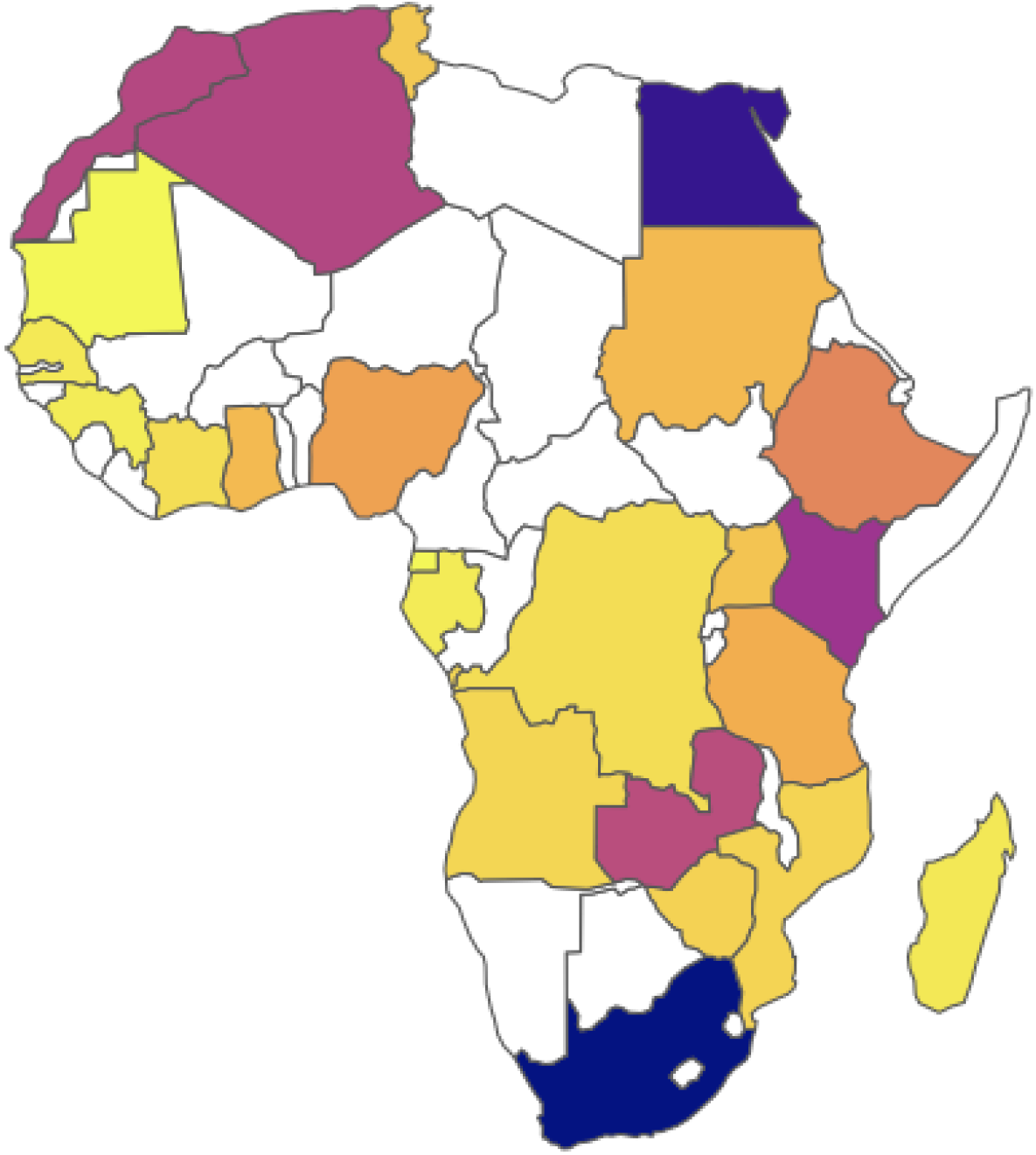
Map of 24 African locations used in the analysis (two locations not shown are the island nations Seychelles and Mauritius) with countries shaded by the magnitude of predicted imported cases from 18 Chinese cities during our focal time period (1st December 2019 to 29th February 2020) under scenario 1. The predicted cases from all scenarios are given in Table S1. The vast majority of predicted cases (100%; 99.9% - 100%) would have occurred prior to any confirmed cases in those locations. Countries shaded in white are locations for which we do not have data for prediction.

**Figure 3.**
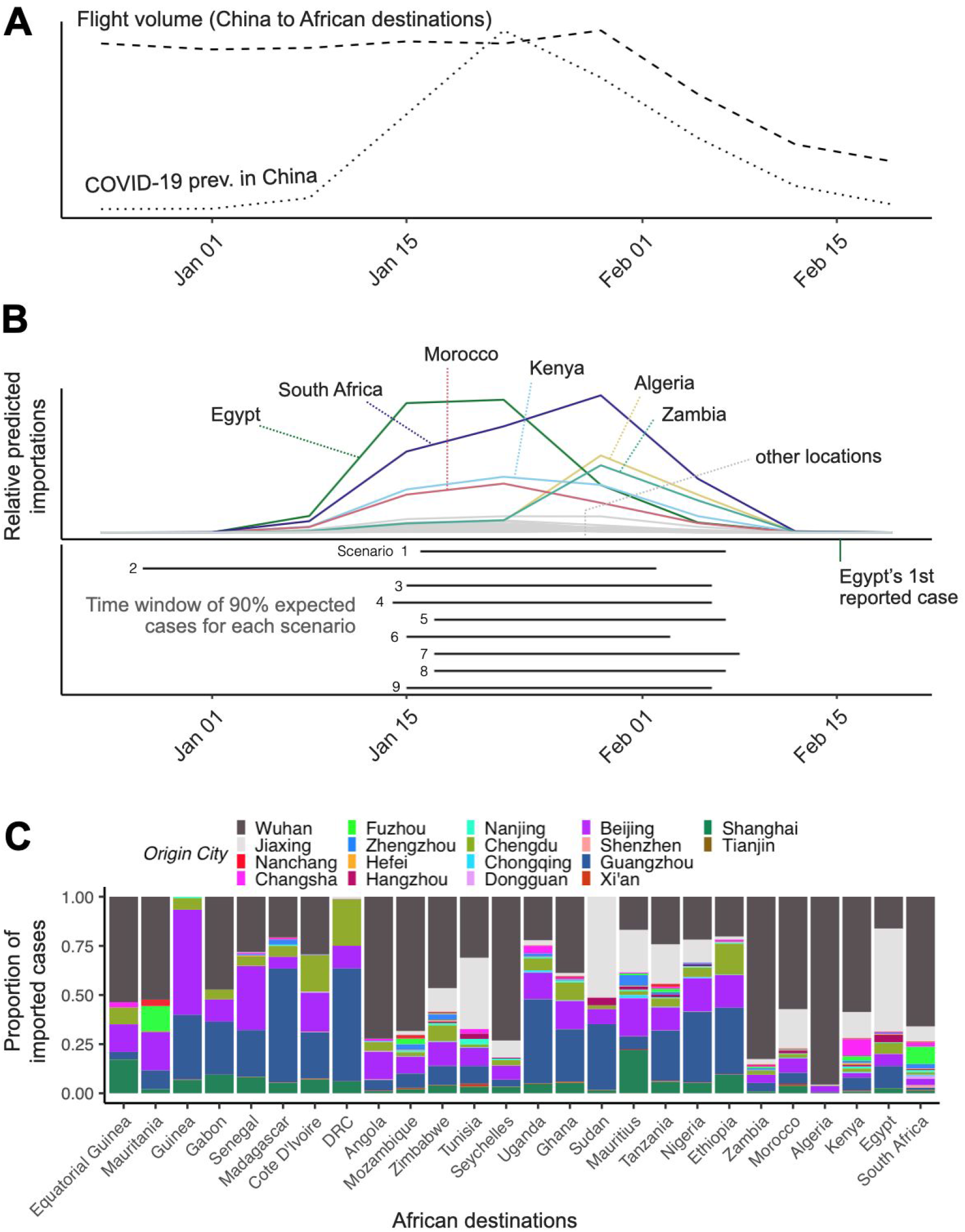
**(A)** Daily prevalence indicator (dotted line) summed across all 18 origin Chinese cities (including Wuhan), from scenario 1, and weekly flight volume from those cities to African destination countries (dashed line) over time. Prevalence peaks on 22nd January 2020, while total flight volume peaks on 29th January 2020. **(B)** Colored and grey curves show weekly predicted number of cases for different destinations in Africa under scenario 1. The first case on the African continent was reported in Egypt, on 15th February^7^ (shown as green vertical line). Our nine model scenarios predict very consistent time windows in which 90% of imported cases are predicted to have arrived, barring scenario 2 (shown as solid horizontal bars; bottom panel). (**C**) Rank of 18 Chinese cities by fraction of all predicted imported COVID-19 cases in each of the 26 countries in Africa included in our analysis, under scenario 1. Countries are ranked from left to right by the total number of imported cases from 1st December 2019 to 29th February 2020. Origin cities are ranked from the bottom to top of each column by maximum estimated prevalence.

### 4. Contribution of different locations in China to globally imported cases

We estimated the relative importance of Wuhan versus other Chinese cities as a source for international case importations and explored how these relative roles changed over time. To do so, we computed the daily forces of importation (as described above in Section 3) to our globally representative locations with two different sources: 1) Wuhan, and 2) the 17 other Chinese cities. We found that early on in the pandemic, the majority of imported cases originated in Wuhan (99%; 58%-100% in the week of 1st January 2020), but this proportion then changed rapidly. The outlying lowest estimate of 58% corresponds to Scenario 2, which predicts substantial prevalence in all provinces in December and early January and higher prevalence in non-Wuhan cities relative to Wuhan compared to the other scenarios. In early January and late February, the proportion of globally imported cases sourced in Wuhan begins to decline non-monotonically, dropping precipitously to 0% in the week of 19th February (across scenarios, Supplementary Figure S4). This indicates a dramatic change in the contribution to imported cases from Wuhan relative to the rest of China. Note that before the contribution of Wuhan is drastically reduced due to the lockdown, one can observe a slight increase in the proportion of cases attributed to Wuhan in late January and early February that can be explained by the rapidly increasing prevalence during a second peak of disease activity (Supplementary Figure S2) during this period.

The contribution of different source populations is expected to further vary across destinations. For the African destinations in our analysis, Wuhan contributed slightly less early in the epidemic (97%; 33% - 98% in the week of 1st January 2020, where 33% corresponds to Scenario 2), subsequently declining in early February to 0% (across scenarios) in mid February (the week of 19th February). Figure 3C further illustrates the variability in the 18 Chinese cities’ contribution to predicted imports across the 26 African destinations. Beijing, Guangzhou, Jiaxing and Chengdu consistently rank among the top cities in terms of their relative contribution to imported cases to each of the African countries included in our analysis. Using the cumulative force of case importation during the early pandemic period from different sources and destinations, we estimated that globally for every one case imported from Wuhan, 0.6 cases (0.1 - 5.6) may have been imported from outside Wuhan. For countries in Africa this ratio was 1.0 (0.1 - 9.8).

### 5. Sensitivity analyses of estimated prevalence indicators

In addition to Scenario 2, our seven other prevalence scenarios tested the robustness of our estimates to uncertainty in three key parameters: 1) ascertainment rates in Hubei relative to non-Hubei provinces; 2) the time that an individual contributed towards the prevalence indicator post-infection; 3) the allocation of province-level case reports to the cities considered here. Varying the ascertainment rate ratio for Hubei to non-Hubei from 2:1 to 1.4:1 (Scenario 3) did not have a significant impact on our results (Table S1). However, we note that our estimates for the absolute number of imported cases were sensitive to the assumed level of under-ascertainment in Hubei versus non-Hubei provinces when we varied the under-ascertainment ratio by one order of magnitude in each direction (Scenarios 6&7). When we assumed that under-ascertainment was 20 times greater in Hubei than outside of Hubei (lower proportion of true cases ascertained in Hubei), the absolute number of predicted importations from cities outside of Hubei dropped to 0.911 cases. Conversely, when assuming that under-ascertainment was 5 times higher outside of Hubei relative to Hubei, the absolute number of predicted imports from cities outside of Hubei increased substantially to 91 cases.

In contrast, varying our assumption for the duration that an infected individual remained infected and eligible for international travel to 2 and 7 days (Scenarios 4&5) did not change our results (Table S1). Although a shorter prevalence duration decreases the magnitude of the prevalence indicator, it has the same impact across Wuhan and non-Wuhan cities and is compensated by the calibration step described in Section 3, so has no impact on the results. We also accounted for the observation that a substantial fraction of cases reported outside of Wuhan originated from Hubei, and were thus only eligible to travel from a non-Wuhan city for the latter stage of their infection, again noting little impact on our results (Scenario 8). Next, rather than assuming that all reported cases were in the cities investigated here, we instead apportioned reported cases to cities equal to their fractional share of that province’s population (Scenario 9, see Methods). This did change the relative contribution of different Chinese cities to total imports – with a noticeably elevated role of Beijing and Shanghai – due to changes in per-capita prevalence. However, overall trends in the number of imported cases over time in the African destination countries remained relatively unchanged, with the exception of Egypt, as did the total number of imports (20.6).

## Discussion

In this study we aimed to make predictions about internationally imported COVID-19 cases from all of China. Our analysis differs from that of previous studies in three fundamental ways: 1) instead of estimating risk of importation,^15^ our model predicted actual number of imported cases, and importantly does so for countries on the African continent; 2) instead of accounting only for travellers from Wuhan,^9^ we accounted for travellers from all major Chinese airports as a potential source population; and 3) we incorporated current air-travel data from December 2019 to February 2020 and back-calculated prevalence from reported COVID-19 cases in China.

These methods in combination enabled us to predict daily time-varying case importations over the first three months of the pandemic. Our model predicted that until the end of February (29 February 2020) 18.4 (range: 8.5 - 100) COVID-19 cases from all of China could have been imported to the 26 African destinations included here. Importantly, our model provided a relatively precise time-frame for those importations. It predicted that the majority (90%) of case importations in these locations occurred between 7th January (±10 day) and 5th February (±3 days). All predicted cases would have been undetected: the first African country to confirm a COVID-19 case was Egypt, which confirmed its first case 14 February 2020^20^, followed by Algeria and Nigeria, with their first confirmed cases on 25 February and 28 February 2020, respectively.^21^ If our predictions are accurate, then undetected imported cases already occurred a month before the first cases were actually confirmed, updating our understanding of the possible timing of when local transmission may have started in those locations. In the absence of strong surveillance systems, estimates of the current and future prevalence rely on dynamic transmission models, with a key unknown being the seeding time of the outbreak.^22,23^ Our results provide, to our knowledge, the first estimates of when index cases may have arrived in different African countries.

We further observed pronounced differences in the number of expected cases imported to the different African destinations. The highest numbers are expected for South Africa, Egypt, Kenya, and Algeria, the lowest numbers for Mauritania, and Equatorial Guinea. These heterogeneities in predicted imports and the relatively early lock-down of borders in several African countries could at least in part explain early differences in the scale of outbreaks in these countries, with some experiencing outbreaks of considerable magnitude and others seemingly spared. Moreover, heterogeneities in the timing of the peaks of predicted importations (Figure 3B) may be due to differences across African countries in changes in air travel volume over the focal time period, or differences in the proportion of passengers flying from Wuhan relative to other Chinese cities over time. On 31st March 2020, a month after the end of our prediction period, South Africa (1,326 cases), Egypt (609 cases), and Algeria (584 cases) were the countries in Africa with the most reported cumulative cases and likely location transmission^24^, while Equatorial Guinea (14 cases) and Mauritania (5 cases) reported relatively few cases,^25^ with unlikely local transmission^24^. Thus, our predictions align with the observed case counts one month later. We expect that as time passes, an increasing number of factors beyond importations from China will influence the observed epidemics in those locations, including sourcing from newly emerging epicenters, differences in response measures, reporting and testing capacity across countries, as well as travel between African countries.

A strength of our method framework is that it rests on a relatively small number of assumptions. For example, it does not rely on estimates of actual case prevalence in China (with the exception of Scenario 2); instead, case counts in high-surveillance countries are used to relate relative force of importation to absolute numbers of cases. It does however need to make assumptions about relative ascertainment rates in Hubei compared to other provinces. To reflect uncertainty in estimates of this ratio, we assumed moderately higher ascertainment rates in other provinces compared to Wuhan based on literature^18^ (Scenarios 1, 3-5), but also the inverse relationship (Scenario 7); we also include a scenario with with ascertainment rates in other provinces strongly exceeding those in Hubei (Scenario 6) and a scenario that reflects time-varying under-reporting (Scenario 2). Additionally, we assumed a fixed value of reporting delays varying over time, although delays were in fact estimated to decrease from late January onwards.^5^ If reporting delays did indeed decline over time, with cases reported from late January^5^ onwards actually incident at a later date than we infer, then this could influence the tail end of the trajectory of predicted imported cases. However, it is unlikely to influence the initial trends in predicted imported cases, the timing of the first introduced cases, or the overall magnitude of imported cases.

Finally, our estimates using daily flight departure data may only approximately reflect the number of passengers flying each day, where more flights might not directly correspond to increased total flight volume and vice versa. This approximation will be increasingly violated in times when demand–and thus airplane capacity–drastically changes, as was probably the case in early 2020. Second, sparsity of recorded flight departures required us to smooth over particularly sparse time periods. However, we expect that these assumptions will have a relatively muted impact on the timing of predicted imporations.

It is important to note that while the magnitude of total predicted imports to the African locations under our best-guess scenarios was relatively modest, this is a direct consequence of the scale of the case data on which we train our model. Specifically, due to low reported case counts in some of our included validation countries, our estimated forces of importation from Wuhan do not require a substantial adjustment to correspond to imported cases in these locations, resulting in forces of importation for all other cities to our African locations that are only marginally adjusted. Furthermore, while we expect the presence of asymptomatics to result in increased predicted imports, by increasing our estimated forces of importation to these locations by an additional factor, we do not expect this to influence the observed shape in the imported case curves over time, if the proportion of asymptomatics among all cases is unchanging over time. Finally, our scenarios capture different epidemic trajectories, in terms of the initial emergence and rate of increase in prevalence, and thus reflect a wide range in possible dynamics of imported cases in the African destination countries. These scenarios all indicate an early timing of initial imported cases, where the scenario assuming a more measured and early increase in prevalence (Scenario 2) places the introduction of the earliest case far before the remaining scenarios.

Recent work has suggested that, globally, at least 62% of cases imported from Wuhan may have remained undetected due to limited surveillance capacity.^6^ Our predictions highlight continued underestimation; when considering all of China as a source of importation–and assuming equivalent sensitivity for detecting cases from Wuhan and other Chinese cities–% (% - %) of all cases imported globally may have been undetected. Wuhan was identified early as the major source population based on its high COVID-19 prevalence. Here, we show the importance of spill-over to locations in the rest of mainland China, and that possible source populations strongly depend on actual travel volume. Some locations may have relatively low SARS-CoV-2 prevalence, but greater connectivity to a given destination country, which would still result in a high overall number of imported infections. Our model predicts for the early pandemic that between 58%-100% of globally imported cases came from Wuhan in the week of 1st January 2020, with the rest originating from other Chinese cities. We find this proportion dropped to 0% in the week of 19th February. This sheds light on how profoundly source populations can change over time under the effect of lockdowns as well as a rapidly spreading virus. In addition, the relative importance of a source population also depends on the destination of imported cases. We found that for the African locations the average proportion of cases exported from Wuhan in the week of 1st January 2020 was slightly lower than that of all destinations, ranging from 33%-98% in the week of 1st January 2020, but similarly declining to 0% in mid-February. This likely reflects differences in business relationships and/or the dominant reasons for travel.

Our findings highlight the importance of a more nuanced understanding of likely sources of case importation for predictive modelling, which will be an important task for ending this pandemic. In sum, this framework allows for routinely quantifying recently imported, and potentially overlooked cases and assessing the principal sources of importation to prioritize in surveillance efforts among travelers, particularly during the later stages of this pandemic when travel restrictions are eased. Going forward, countries wishing to identify likely sources of case importations may benefit from combining ongoing travel volume and prevalence data as we have done here, allowing for more nuanced policy decisions than those based on global trends. Our approach further aids in elucidating the timing of the first imported cases, which is crucial for initializing models aimed at anticipating future trends in COVID-19 transmission in diverse locations. Importantly, these methods can be adjusted to incorporate prevalence estimates and flight data from any number of origin and destination locations with minimal data requirements: reported case data in the source and calibration locations, daily flight departure data and monthly flight passenger totals. The tools we propose here are particularly useful for locations facing significant surveillance constraints, and potential resource limitations in managing ongoing response efforts, and thereby work to address enduring gaps in infectious disease monitoring and preparedness.

## Methods

### 1. Estimating number of airplane passengers from 18 Chinese cities to international destinations

#### Data

We used data on the number of passengers and flight departures from 1st December 2019 to 29th February 2020 from 18 Chinese origin cities to 43 international destinations, as described below. Historical air-travel volume data are likely not representative of the pandemic time period for two reasons: 1) Lunar New Year was earlier than in preceding years (25th January 2020), and 2) large-scale travel bans and flight cancellations took place in late January 2020.

We defined three main categories of locations as international destinations in our analyses 1) ten international locations with high surveillance capacity and high air travel connectivity to Wuhan used for model calibration; 2) 26 African countries as destinations used for model prediction; and 3) 16 locations that represent the top three destinations (two for Oceania and North America) from each inhabited continent (Oceania, Asia, Africa, North America, South America) in terms of travel volume from China during our focal time period. Those 16 locations include locations from 1) and 2) and an additional seven locations. We used these locations to estimate the ratio of cases imported internationally from other Chinese origin cities compared to cases imported internationally from Wuhan. For all flight data, in addition to Wuhan, we included as origin locations (*N*_*O*_ =17) the 17 Chinese cities that were previously identified by Lai *et al*. (2020)^11^ as high-risk cities for importation of COVID-19 from Wuhan and therefore likely sources of imported cases internationally: Beijing, Shanghai, Guangzhou, Zhengzhou, Tianjin, Hangzhou, Jiaxing, Changsha, Nanjing, Nanchang, Shenzhen, Chongqing, Chengdu, Hefei, Fuzhou, Xi’an, and Donngguan.

For destinations outside of China, we considered a selection of locations with both high air travel connectivity to Wuhan and high surveillance capacity (thus “high surveillance locations”) for model validation. We assessed surveillance capacity using the Global Health Security (GHS) Index, in particular its components of “early detection and reporting epidemics of potential international concern”, published in 2019.^6^ We thus selected locations with the highest connectivity to Wuhan as estimated by Lai *et al*. (2020)^11^, and within the top 5% percentile of the GHS index rank. We additionally included Singapore as it has demonstrated a strong capacity to identify, trace and document COVID-19 cases^9,26^, despite having a relatively low GHS index.

In total, we used *N*_*D*_ = 43 total destination locations: 1) the ten high-surveillance locations of Singapore, US, Australia, Canada, South Korea, UK, Netherlands, Sweden, Germany, and Spain for model validation; 2) 26 African countries for prediction, representing the 26 top destination cities in Africa in terms of air-travel volume from 18 high-risk cities in mainland China (Nigeria, Ghana, Algeria, Côte D’Ivoire, Ethiopia, Egypt, Guinea, Morocco, Tanzania, Senegal, South Africa, Uganda, Congo (Kinshasa), Zimbabwe, Sudan, Angola, Gabon, Zambia, Mozambique, Mauritania, Mauritius, Kenya, Seychelles, Madagascar, Tunisia, Equatorial Guinea); and 3) 16 global main destinations from China (New Zealand, Australia, United Kingdom, Germany, Russia, Japan, Thailand, Korea (South), United States, Canada, Brazil, Argentina, Chile, Egypt, Ethiopia, South Africa).

We used data from International Air Transport Association (IATA)^27^ on the monthly number of confirmed passengers on flights (direct and indirect) for each of the *N* origin-destination pairs, henceforth referred to as air-travel volume, from December 2019 to February 2020. In addition to air-travel volume, we used data from Cirium^28^ on the number of daily departed (and landed) direct passenger flights for each of the *N* origin-destination pairs, henceforth referred to as ‘flight departures’, for the period December 2019 to February 2020.

We combined the Cirium data on daily flight departures with the IATA monthly air-travel volume data to estimate daily air-travel volume out of cities in China to our destinations of interest from 1st December 2019 to 29th February 2020. For all origins except for Wuhan, we distributed the monthly air-travel volume in month *m, V* _*i,j,m*_, into daily air-travel volume for each day *d*, using the proportion of daily flight departures out of the total number of daily flights in the corresponding month.

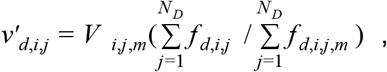

where *v* ′_*d,i,j*_ is the imputed air-travel volume from origin *i* to destination *j* on day *d, f* _*d,i,j,m*_ is the number of landed flight departures on day *d* from origin *i* to destination *j* in month *m*, and *N*_*D*_ is the number of destination locations. When distributing the monthly air-travel volume, we used the proportion of daily flight departures for each origin *i* summed over all destinations *j* instead of calculating this for each origin-destination connection due to sparse data limitations. Furthermore, given that flight departures to each of the 6 continents in our analysis exhibited similar trends over time (results not shown), we chose not to disaggregate this measure within a continent.

The same approach could not be applied for Wuhan due to sparsity in the number of direct landed flight departures from Wuhan in the Cirium data after 23rd January 2020. For Wuhan, to distribute monthly air-travel volume, *V* _*i,j,m*_, into daily air-travel volume for each day, we instead fit a smoothing spline to the daily number of landed flight departures and used its predictions as inputs in the above equation. We used the “forward-chaining” time-series cross-validation procedure, applying an accumulative rolling training window of seven days, in order to estimate the smoothing parameter for the spline.^29,30^

Since residents of the cities Shenzhen and Dongguan both use the Shenzhen Bao’an International Airport, we divided the air-travel volume equally between these two cities. Similarly, the air-travel volume for Hangzhou Xiaoshan International Airport airport is divided equally for the cities Jiaxing and Hangzhou. Importantly, Cirium data only documents the daily number of direct flights, but we use this data source to distribute a total three-month volume into daily volume. We therefore make the assumption that the variation over time in the number of direct flights is proportional to the variation in the number of direct and indirect flights.

### 2. Estimating daily prevalence of COVID-19 in 18 Chinese cities

We used data on the number of confirmed COVID-19 cases reported by China CDC per day by province^31,32^.To estimate daily incidence, we backwards shifted the time series of confirmed cases by a mean reporting delay of seven days^5^ to yield the total number of symptom onsets per day. We shifted this onset incidence curve backwards again by a median incubation period of five days^17^ to yield the total number of infection onsets who had not yet developed symptoms. The mean reporting delay is estimated using line list data summarized in Zhang et al. 2020, which provides information on the number of days after which an individual develops symptoms when someone is reported as a case^5^. We also tested a more accurate back-calculation method for estimating infection incidence by deconvoluting the case confirmation curve and the time-varying reporting delay distribution (derived from the line list data). Exploratory analysis using simulated data revealed that this additional complexity had a negligible impact on inferred infection incidence (results not shown).

We then scaled infection onset curves by province by our assumed ascertainment rate ratio, which defines the ascertainment rate in all other provinces relative to that of Hubei (e.g. an ascertainment rate ratio of 2:1 implies that twice as many true cases were reported in non-Hubei provinces relative to Hubei). Estimates of this ascertainment rate ratio are derived from (1) Verity et al.^18^ and (2) from Maier and Brockman^33^. For (1), we evaluated the relative reporting rate using the digitized estimates of ascertainment rates across nine age groups–with an assumed referent group of the 50-59 years age group outside of Wuhan–stratified by Wuhan and cities outside of Wuhan^18^. To do so, we calculated the ratio of the sum of population-weighted estimates of ascertainment rates by age group for cities outside of Wuhan to the sum of population-weighted estimates of the ascertainment rates by age group for Wuhan. For (2), we evaluated the relative reporting rate using digitized plots of “unidentified infecteds” and confirmed cases from January 21st to March 1st in Hubei and in all other provinces (combined)^33^. We defined ascertainment rates separately for Hubei and for all other provinces as one minus the proportion of total cases that were “unidentified infected”, estimated at two instances before and after an inflection point marking the point when confirmed cases eclipse “unidentified infecteds”^33^. We select times during which the differential between the confirmed case and “unidentified infected” curves is approximately maximized to yield the lowest possible and highest possible ascertainment rates in these two periods^33^. The overall ascertainment rate over the entire period is then estimated by averaging the two period-specific ascertainment rates, weighted by the proportion of days they contribute, separately for Hubei and for all other provinces. Finally, we took the ratio of the overall ascertainment rates in all other provinces and in Hubei to estimate the ascertainment rate ratio. We selected the Verity et al. estimate of 2:1 as our best guess value. As a further sensitivity analysis, we varied this ratio by one order of magnitude in each direction to give 0.2:1 and 20:1.

As an alternative to our back-shifted incidence curves, we also used estimated onset incidence curves from a previous Bayesian analysis of SARS-CoV-2 onset incidence in China^19^. Tsang et al. make the crucial point that case ascertainment driven by changes in case definition over time would have a significant impact on the inferred dynamics of the epidemic. The authors used a Bayesian analysis assuming exponential growth (with a different rate before and after 23rd January 2020) to infer the number of case onsets that would have been observed had a later, broader COVID-19 case definition been used throughout the outbreak. For Wuhan, we used the posterior mean estimated onset incidence curve (Figure 3 of Tsang et al.^19^), which estimates onset incidence in Wuhan if the COVID-19 case definition as of 4th February 2020 had been applied throughout. For non-Wuhan cities, we took the posterior mean estimated onset incidence curve for China excluding Wuhan divided among each Chinese province proportional to the number of confirmed cases in the province from China CDC data^31,32^. Finally, we back-shifted these onset curves by the median incubation period as above.

For all scenarios, we estimated the travel-related prevalence indicator (corresponding to absolute prevalence in the case of scenario 2) each day by summing over individuals who were infected on a given day and individuals who were infected in previous days and have not yet developed symptoms (Table 1). To convert our estimates of the province-level prevalence indicator to per-capita prevalence in each Chinese city, we divided all prevalent infections in each province (absolute numbers) equally across all of the cities that we consider in our analysis that are in that province. In so doing, we assume that individuals in a province are equally likely to go to the specific airports in our analysis. For example, we assumed that 100% of cases in Jiangxi province were in Nanchang with a population of around 2.4 million, whereas we assumed that one third of cases in Guangdong were in Guangzhou, Dongguan and Shenzhen respectively. As a sensitivity analysis, Scenario 9 instead assumed that prevalent cases were attributed to cities proportional to the percentage of the population in that city. For example, the city of Hefei accounts for 5.23% of the population of Anhui, and was therefore assumed to account for 5.23% of all infections in Anhui. Finally, we divided the total number of cases by the population size of that city to generate per-capita prevalence estimates.

### 3. Estimating number of imported cases to international destinations

#### 3.1 Model training: Associating flight volume of infected passengers from Wuhan with observed number of Wuhan-origin cases in validation set locations

We first fit a model to the number of imported COVID-19 cases from Wuhan observed in the high surveillance locations to determine the relationship between prevalence, air-travel volume and imported case counts. We used this model fit to make predictions using data from Wuhan and the remaining cities in China. The number of observed cumulative cases imported from Wuhan to destination *j* in prior to the 23rd January is denoted as *y*_*j*_. Further, 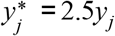 denotes the number of cases that each destination location *j*, excluding Singapore, could have detected with a surveillance capacity of Singapore^6^ (for Singapore 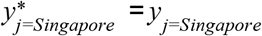). Following the analysis by Niehus et al., 2.5 represents the ratio of Singapore’s “capacity” to identify imported cases to that of other high-surveillance countries.^6^ We assumed that across the high surveillance destinations (U.S., Australia, Canada, South Korea, UK, Netherlands, Sweden, Germany, Spain, and Singapore), this number follows a Poisson distribution, as follows:

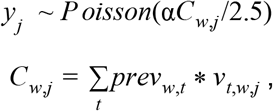

where *C*_*w,j*_ represents the force of importation from Wuhan to each destination *j*, which is calculated as the product of the COVID-19 prevalence indicator in Wuhan (*prev*_*w,t*_) and volume of passengers from Wuhan to destination *j* (*v*_*t,w,j*_) on day *t*, summed over all days in the pre-lockdown pandemic period, and α represents the scaling factor relating the force of importation to scaled reported cases in the high surveillance locations,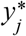. We fit this model using the *glm* function in R (version 3.6.1).^34^

### 3.2 Model application: Predicting imported case counts to subset of African destinations

We defined the pre-lockdown pandemic period (referring to the lockdown of Wuhan) and our focal pandemic period, which we considered to be from 1st December 2019 (approximate date of seeding in Wuhan^15^) to 23rd January 2020 and from 1st December 2019 to 29th February 2020, respectively.

The force of importation of COVID-19 from all selected cities in China to destination *j* in Africa was computed as:

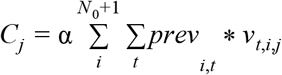

where *prev*_*i,t*_ is the prevalence indicator of COVID-19 in Chinese city *i* at time *t*, and *v*_*t,i,j*_ is the total volume of passengers across flights from each origin city *i* to each destination *j* at time *t*. The product of daily passenger volume (*v*_*t,i,j*_) and the COVID-19 prevalence indicator in city *i* (*prev*_*i,t*_) was summed over all days of the focal pandemic time period, and over all *N*_*0*_ Chinese cities and Wuhan. We used this force of importation to make predictions for all 26 African locations under each of the five scenarios for the *N*_*0*_ Chinese cities.

### 3.3 Ratio of force of importation from Wuhan compared to the rest of China

To estimate the ratio of expected imported cases from Chinese cities without Wuhan versus expected imported cases from Wuhan, we computed both the force of importation from Wuhan (*C*_*w,j*_, as defined above) and the force of importation from the 17 Chinese cities, excluding Wuhan, as follows:

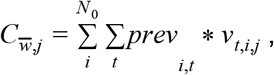

where the product of daily passenger volume (*v*_*t,i,j*_) and the COVID-19 prevalence indicator in city *i* (*prev*_*i,t*_) under Scenario 1 is summed over all days of the focal pandemic period, and over the 17 Chinese cities (excluding Wuhan). The ratio of 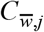 and *C*_*w,j*_ gives the ratio of expected cases exported from outside Wuhan and cases exported from Wuhan. We computed this ratio for two different sets of locations (*j*∈{African locations} and *j*∈{all locations}), under scenario 1, according to:

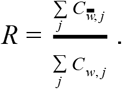

We repeated this procedure with the remaining eight prevalence scenarios to bound our estimates of *R*.

## Data Availability

Transformed flight data, derived prevalence data, and all code used in these analyses to be made fully available online at https://github.com/c2-d2/COVID_allchina_export

## Supporting Information

**Supplementary Figure S1.**
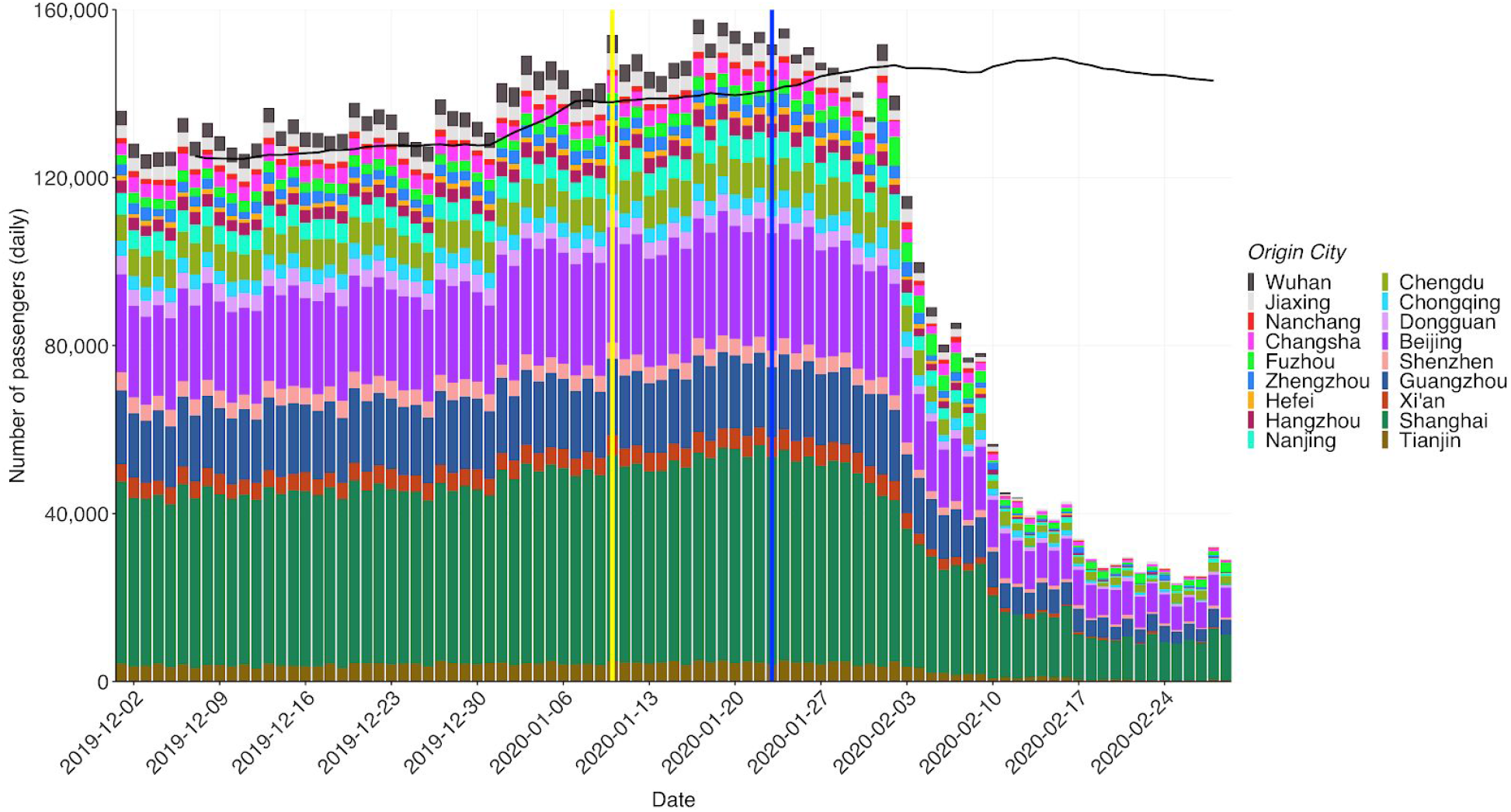
Estimated daily flight volume from 18 Chinese cities to 43 international destinations (10 high-surveillance destinations for model validation, 26 African destinations, 7 additional destinations) from 1st December 2019 to 29th February 2020. The yellow vertical line marks the start of the 40-day Chunyun period surrounding Lunar New Year. The blue vertical line indicates 23rd January 2020, the day travel restrictions were widely implemented in Wuhan. The black line shows the 7-day rolling average of the estimated daily flight volume from 18 Chinese cities to 43 international destinations for the equivalent period one year earlier: from 1st December 2018 to 28th February 2019.

**Supplementary Figure S2.**
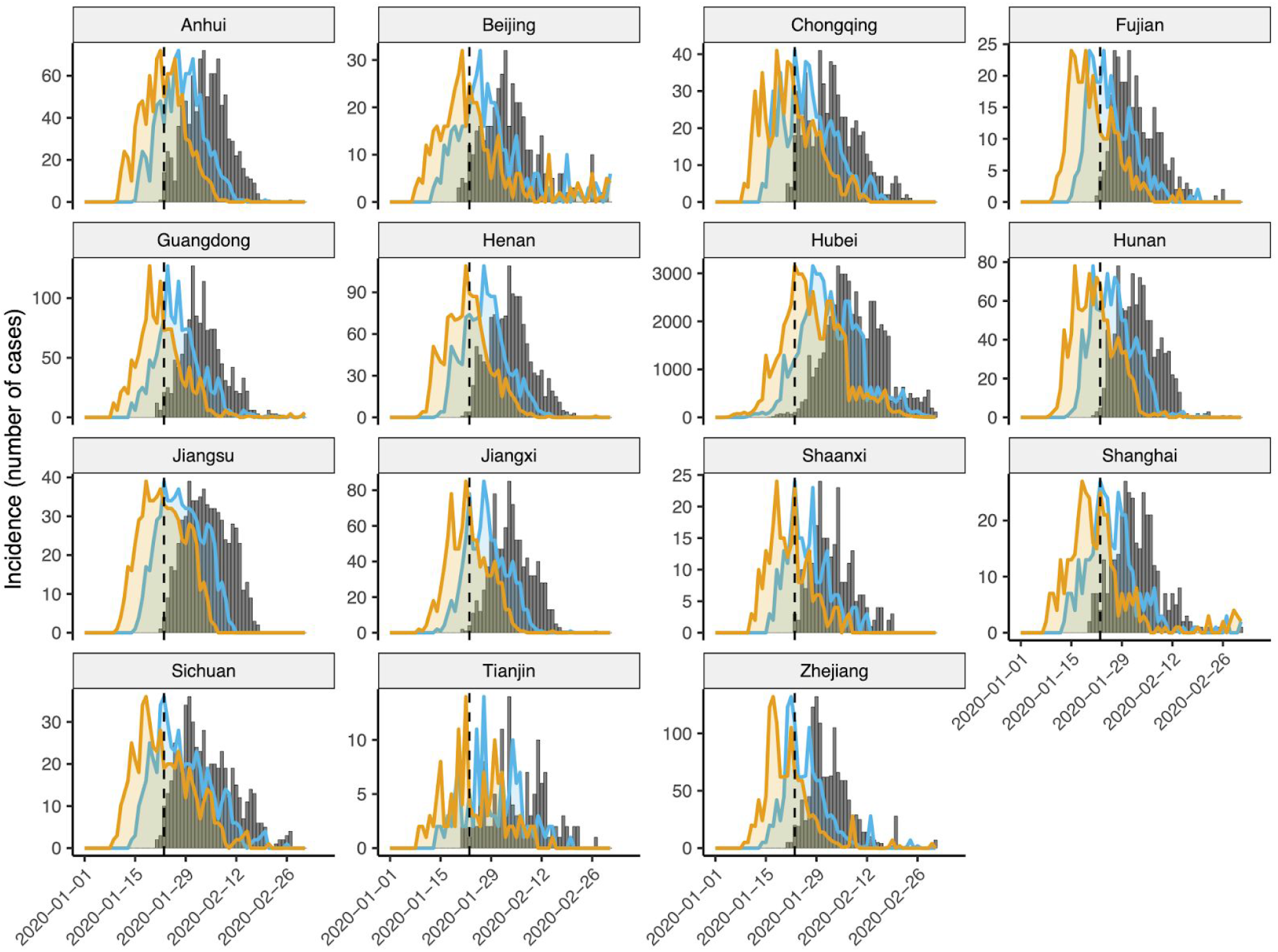
Back-calculation of symptom onsets (blue) and infections (orange) from observed confirmed case counts (grey bars) for all provinces considered here. Confirmed cases were shifted back by 7 days (the mean confirmation delay) to estimate symptom onset incidence, then further by 5 days (the median incubation period) to estimate infection incidence of those cases. Vertical dashed line shows 23rd January 2020, the date of lockdown in Wuhan.

**Supplementary Figure S3.**
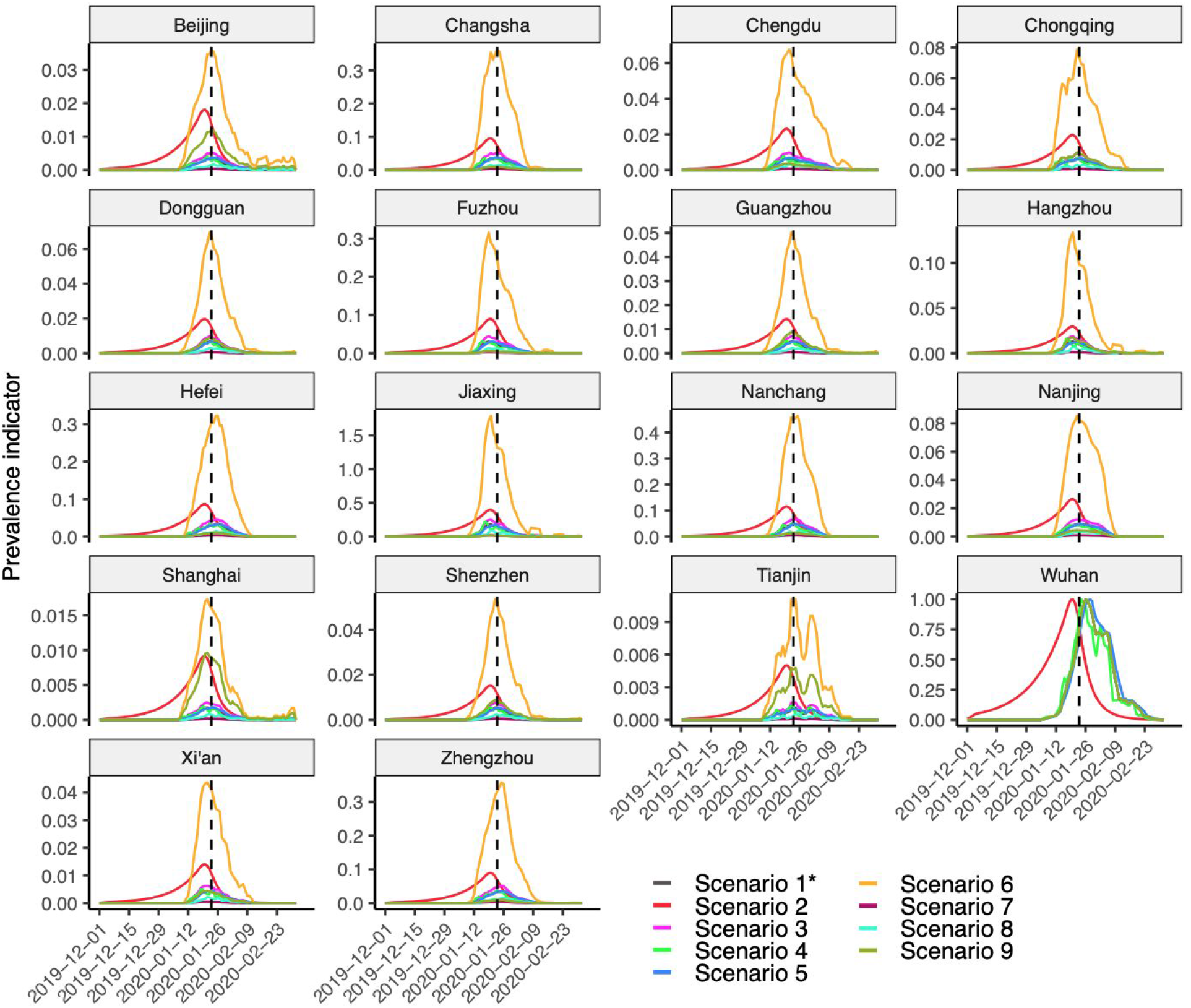
Prevalence indicator in the 18 considered Chinese cities. Values were scaled relative to the maximum prevalence indicator in Wuhan within each scenario. Note that Scenarios 1, 3 and 6-9 yield identical curves in Wuhan. Note also that Scenario 2 is based on independent estimates as described in the main text Methods. Vertical dashed line shows 23rd January 2020, the date of lockdown in Wuhan.

**Supplementary Figure S4.**
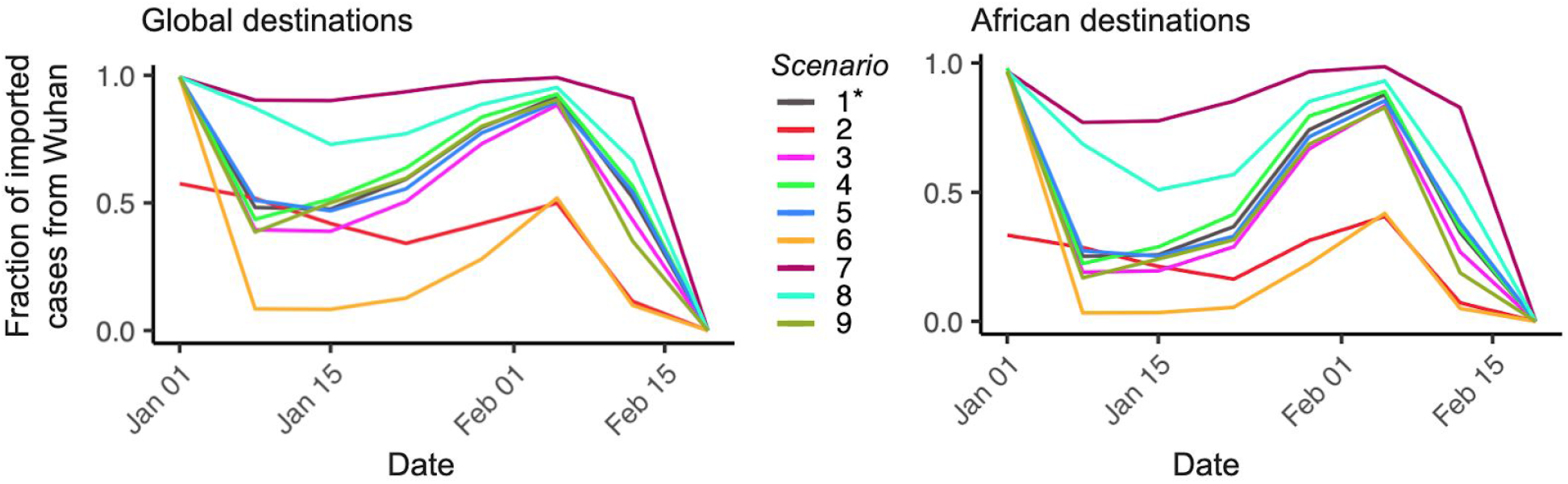
Weekly fraction of imported cases from Wuhan to global destinations (left-hand side) and to African destinations (right-hand side) shows as mean predictions for each of the 9 scenarios. The best-estimate scenario is Scenario 1 (highlighted with an asterisk). The x-axis shows the date for the current year 2020.

**Supplementary Table S1.** Table of mean imported cases to 26 African destination locations, by scenario, with our best-guess scenarios marked by a *. Countries ordered alphabetically.

| Country | 1* | 2 | 3 | 4 | 5 | 6 | 7 | 8 | 9 |
| --- | --- | --- | --- | --- | --- | --- | --- | --- | --- |
| Algeria | 1.56 | 0.21 | 1.59 | 1.15 | 1.92 | 2.18 | 1.50 | 1.52 | 1.68 |
| Angola | 0.13 | 0.07 | 0.14 | 0.11 | 0.15 | 0.45 | 0.10 | 0.11 | 0.18 |
| DRC | 0.10 | 0.07 | 0.15 | 0.07 | 0.12 | 1.02 | 0.01 | 0.04 | 0.19 |
| Côte D'Ivoire | 0.10 | 0.07 | 0.13 | 0.08 | 0.12 | 0.74 | 0.04 | 0.06 | 0.19 |
| Egypt | 3.89 | 2.45 | 5.29 | 3.02 | 4.54 | 33.23 | 0.95 | 1.91 | 3.29 |
| Equatorial Guinea | 0.04 | 0.02 | 0.04 | 0.03 | 0.04 | 0.19 | 0.02 | 0.03 | 0.08 |
| Ethiopia | 0.63 | 0.40 | 0.84 | 0.47 | 0.76 | 5.15 | 0.18 | 0.33 | 1.24 |
| Gabon | 0.06 | 0.04 | 0.08 | 0.05 | 0.07 | 0.36 | 0.03 | 0.04 | 0.12 |
| Ghana | 0.24 | 0.15 | 0.30 | 0.19 | 0.28 | 1.57 | 0.11 | 0.15 | 0.41 |
| Guinea | 0.06 | 0.06 | 0.08 | 0.04 | 0.07 | 0.59 | 0.01 | 0.02 | 0.16 |
| Kenya | 1.95 | 0.84 | 2.30 | 1.53 | 2.28 | 9.21 | 1.22 | 1.46 | 1.79 |
| Madagascar | 0.07 | 0.05 | 0.10 | 0.06 | 0.08 | 0.58 | 0.02 | 0.04 | 0.13 |
| Mauritania | 0.04 | 0.02 | 0.04 | 0.03 | 0.05 | 0.20 | 0.02 | 0.03 | 0.06 |
| Mauritius | 0.28 | 0.20 | 0.37 | 0.21 | 0.32 | 2.34 | 0.07 | 0.14 | 0.62 |
| Morocco | 1.52 | 0.77 | 1.80 | 1.21 | 1.75 | 7.38 | 0.93 | 1.12 | 1.82 |
| Mozambique | 0.14 | 0.06 | 0.15 | 0.10 | 0.17 | 0.52 | 0.10 | 0.11 | 0.17 |
| Nigeria | 0.38 | 0.26 | 0.51 | 0.29 | 0.45 | 3.07 | 0.11 | 0.20 | 0.68 |
| Senegal | 0.07 | 0.05 | 0.09 | 0.05 | 0.08 | 0.52 | 0.02 | 0.04 | 0.16 |
| Seychelles | 0.18 | 0.08 | 0.21 | 0.15 | 0.21 | 0.63 | 0.14 | 0.15 | 0.23 |
| South Africa | 4.51 | 1.72 | 5.17 | 3.47 | 5.37 | 18.34 | 3.13 | 3.60 | 4.41 |
| Sudan | 0.24 | 0.16 | 0.35 | 0.19 | 0.28 | 2.42 | 0.02 | 0.09 | 0.25 |
| Tanzania | 0.31 | 0.20 | 0.40 | 0.24 | 0.36 | 2.39 | 0.10 | 0.17 | 0.46 |
| Tunisia | 0.17 | 0.10 | 0.22 | 0.13 | 0.21 | 1.25 | 0.07 | 0.10 | 0.19 |
| Uganda | 0.20 | 0.13 | 0.27 | 0.15 | 0.24 | 1.60 | 0.06 | 0.11 | 0.35 |
| Zambia | 1.42 | 0.25 | 1.52 | 1.03 | 1.75 | 3.64 | 1.19 | 1.27 | 1.58 |
| Zimbabwe | 0.14 | 0.09 | 0.18 | 0.12 | 0.16 | 0.83 | 0.07 | 0.10 | 0.19 |
| **Total** | **18.4** | **8.5** | **22.3** | **14.2** | **21.8** | **100** | **10.2** | **12.9** | **20.6** |

## Contributors

TFM, TC, JH, PMDS, ML, and RN designed the study. TFM, TC, JH, AWCY, and RN performed the analyses. TFM, TC, JH, PMDS, and RN wrote the manuscript. All authors reviewed and approved the final manuscript.

## Data availability

Transformed flight data, derived prevalence data, and all code used in these analyses are fully available online at https://github.com/c2-d2/COVID_allchina_export.

## Acknowledgments

We thank Mauricio Santillana, Lee Kennedy-Shaffer, Rebecca Kahn, Christine Tedijanto, Justin Lessler, Kylie Ainslie and Charlie Whittaker for their valuable input and feedback. ML, TFM, TC, JAH, MJM and RN were supported by Award Number U54GM088558 from the US National Institute Of General Medical Sciences. PMD was supported by the Fellowship Foundation Ramon Areces. COB was supported by a NIGMS Maximizing Investigator’s Research Award (MIRA) R35GM124715-02. MJM was supported by an NIH DP5 grant.

The content is solely the responsibility of the authors and does not necessarily represent the official views of the National Institute Of General Medical Sciences or the National Institutes of Health.

## References

1. Zhou P, Yang X-L, Wang X-G, et al. A pneumonia outbreak associated with a new coronavirus of probable bat origin. Nature 2020;579(7798):270–3.

2. Lai S, Ruktanonchai NW, Zhou L, et al. Effect of non-pharmaceutical interventions to contain COVID-19 in China. Nature 2020;1–7.

3. Tian H, Liu Y, Li Y, et al. An investigation of transmission control measures during the first 50 days of the COVID-19 epidemic in China. Science [Internet] 2020 [cited 2020 Apr 20];Available from: https://science.sciencemag.org/content/early/2020/03/30/science.abb6105

4. Kraemer MUG, Yang C-H, Gutierrez B, et al. The effect of human mobility and control measures on the COVID-19 epidemic in China. Science [Internet] 2020 [cited 2020 Apr 20];Available from: https://science.sciencemag.org/content/early/2020/03/25/science.abb4218

5. Zhang J, Litvinova M, Wang W, et al. Evolving epidemiology and transmission dynamics of coronavirus disease 2019 outside Hubei province, China: a descriptive and modelling study. Lancet Infect Dis [Internet] 2020 [cited 2020 Apr 20];0(0). Available from: https://www.thelancet.com/journals/laninf/article/PIIS1473-3099(20)30230-9/abstract

6. Niehus R, De Salazar PM, Taylor AR, Lipsitch M. Using observational data to quantify bias of traveller-derived COVID-19 prevalence estimates in Wuhan, China. Lancet Infect Dis 2020;S1473309920302292.

7. ECDC. opendata ECDC [Internet]. [cited 2020 May 20];Available from: https://opendata.ecdc.europa.eu/covid19/casedistribution/csv

8. Clinical management of severe acute respiratory infection when novel coronavirus (2019-nCoV) infection is suspected: Interim guidance [Internet]. World Health Organization; 2020. Available from: https://www.who.int/docs/default-source/coronaviruse/clinical-management-of-novel-cov.pdf

9. Salazar PMD, Niehus R, Taylor A, Buckee CO, Lipsitch M. Identifying Locations with Possible Undetected Imported Severe Acute Respiratory Syndrome Coronavirus 2 Cases by Using Importation Predictions. Emerg Infect Dis [Internet] 2020 [cited 2020 May 24];26(7). Available from: https://www.nc.cdc.gov/eid/article/26/7/20-0250_article

10. Du Z, Wang L, Cauchemez S, et al. Risk for Transportation of Coronavirus Disease from Wuhan to Other Cities in China. Emerg Infect Dis [Internet] 2020 [cited 2020 May 24];26(5). Available from: https://www.nc.cdc.gov/eid/article/26/5/20-0146_article

11. Lai S, Bogoch I, Ruktanonchai N, et al. Assessing spread risk of Wuhan novel coronavirus within and beyond China, January-April 2020: a travel network-based modelling study. medRxiv 2020;2020.02.04.20020479.

12. World Health Organization, Regional Office for Africa. COVID-19 Situation update for the WHO African Region: External Situation Report 2 [Internet]. WHO, Regional Office for Africa; 2020. Available from: https://apps.who.int/iris/bitstream/handle/10665/331425/SITREP_COVID-19_WHOAFRO_20200311-eng.pdf

13. WHO, Regional Office for Africa. More than 15 countries in Africa report COVID-19 cases [Internet]. WHO Reg. Off. Afr. 2020 [cited 2020 Mar 16];Available from: https://www.afro.who.int/news/more-15-countries-africa-report-covid-19-cases

14. Senghore M, Savi MK, Gnangnon B, Hanage WP, Okeke IN. Leveraging Africa’s preparedness towards the next phase of the COVID-19 pandemic. Lancet Glob Health 2020;S2214109X20302345.

15. Gilbert M, Pullano G, Pinotti F, et al. Preparedness and vulnerability of African countries against importations of COVID-19: a modelling study. The Lancet [Internet] 2020 [cited 2020 Feb 27];0(0). Available from: https://www.thelancet.com/journals/lancet/article/PIIS0140-6736(20)30411-6/abstract

16. Volz E, Baguelin M, Bhatia S, et al. Report 5: Phylogenetic analysis of SARS-CoV-2 [Internet]. Imperial College London; 2020. Available from: https://www.imperial.ac.uk/media/imperial-college/medicine/sph/ide/gida-fellowships/Imperial-College-COVID19-phylogenetics-15-02-2020.pdf

17. Lauer SA, Grantz KH, Bi Q, et al. The Incubation Period of Coronavirus Disease 2019 (COVID-19) From Publicly Reported Confirmed Cases: Estimation and Application. Ann Intern Med 2020;172(9):577–82.

18. Verity R, Okell LC, Dorigatti I, et al. Estimates of the severity of coronavirus disease 2019: a model-based analysis. Lancet Infect Dis [Internet] 2020 [cited 2020 May 24];0(0). Available from: https://www.thelancet.com/journals/laninf/article/PIIS1473-3099(20)30243-7/abstract

19. Tsang TK, Wu P, Lin Y, Lau EHY, Leung GM, Cowling BJ. Effect of changing case definitions for COVID-19 on the epidemic curve and transmission parameters in mainland China: a modelling study. Lancet Public Health 2020;5(5):e289–96.

20. World Health Organization, Regional Office for the Eastern Mediterranean. Update on COVID-19 in the Eastern Mediterranean Region [Internet]. World Health Organ. 2020 [cited 2020 Mar 20];Available from: http://www.emro.who.int/media/news/update-on-covid-19-in-the-eastern-mediterranean-region.html

21. World Health Organization, Regional Office for Africa. COVID-19 Situation update for the WHO African Region: External Situation Report 1 [Internet]. WHO, Regional Office for Africa; 2020. Available from: https://apps.who.int/iris/bitstream/handle/10665/331330/SITREP_COVID-19_WHOAFRO_2 0200304-eng.pdf

22. Lourenco J, Paton R, Ghafari M, et al. Fundamental principles of epidemic spread highlight the immediate need for large-scale serological surveys to assess the stage of the SARS-CoV-2 epidemic [Internet]. Epidemiology; 2020 [cited 2020 May 21]. Available from: http://medrxiv.org/lookup/doi/10.1101/2020.03.24.20042291

23. Altarelli F, Braunstein A, Dall’Asta L, Lage-Castellanos A, Zecchina R. Bayesian Inference of Epidemics on Networks via Belief Propagation. Phys Rev Lett 2014;112(11):118701.

24. World Health Organization. Coronavirus disease 2019 (COVID-19) Situation Report – 67 [Internet]. World Health Organ. 2020 [cited 2020 Jul 7];Available from: https://www.who.int/docs/default-source/coronaviruse/situation-reports/20200327-sitrep-67-covid-19.pdf?sfvrsn=b65f68eb_4

25. European Centre for Disease Prevention and Control. Situation update – worldwide [Internet]. Eur. Cent. Dis. Prev. Control. 2020 [cited 2020 Feb 21];Available from: https://www.ecdc.europa.eu/en/geographical-distribution-2019-ncov-cases

26. Pung R, Chiew CJ, Young BE, et al. Investigation of three clusters of COVID-19 in Singapore: implications for surveillance and response measures. The Lancet [Internet] 2020 [cited 2020 Mar 24];0(0). Available from: https://www.thelancet.com/journals/lancet/article/PIIS0140-6736(20)30528-6/abstract

27. BlueDot. BlueDot [Internet]. [cited 2020 Mar 10];Available from: https://bluedot.global

28. Cirium homepage [Internet]. [cited 2020 Mar 10];Available from: https://www.cirium.com

29. Hyndman RJ. Cross-validation for time series [Internet]. Hyndsight Blog. 2016 [cited 2020 May 7];Available from: https://robjhyndman.com/hyndsight/tscv/

30. Cochrane C. Time Series Nested Cross-Validation [Internet]. Data Sci. 2018 [cited 2020 May 24];Available from: https://towardsdatascience.com/time-series-nested-cross-validation-76adba623eb9

31. China CDC. Distribution of new coronavirus pneumonia [Internet]. [cited 2020 Apr 1];Available from: http://2019ncov.chinacdc.cn/2019-nCoV/

32. MIDAS Online Portal for COVID-19 Modeling Research [Internet]. [cited 2020 Apr 1];Available from: https://midasnetwork.us/covid-19/

33. Maier BF, Brockmann D. Effective containment explains subexponential growth in recent confirmed COVID-19 cases in China. Science 2020;368(6492):742–6.

34. {R Development Core Team, R}. R: A Language and Environment for Statistical Computing. Available from: http://dx.doi.org/10.1007/978-3-540-74686-7

